# Applying Diagnostic Network Optimisation analysis to inform the introduction of G6PD testing into Indonesia for improved treatment of relapsing malaria

**DOI:** 10.64898/2026.11.03.26367074

**Authors:** Karina Dian Lestari, Ihsan Fadilah, Illene Nanine, Hellen Dewi Prameswari, Minerva Theodora, Bayu Kurnia, Hermawan Susanto, Ratih Ketana Hapsari, Dimas Prayoga, Aditya Kris Nugroho, Made Bhagaskara Hadi Prawira, Riskha Tiara Puspa Dewi, Herdiana, Iqbal R F Elyazar, Rosalind Howes, Ari Winasti Satyagraha

## Abstract

**Background:** Radical cure for *P. vivax* and *P. ovale* malaria using 8-aminoquinoline drugs primaquine or tafenoquine can prevent relapse, but requires glucose-6-phosphate dehydrogenase (G6PD) screening due to the risk of haemolysis in G6PD-deficient individuals. This study aimed to provide an evidence-based framework to inform the optimal allocation and referral networks for G6PD testing devices in Indonesia.

**Methods:** A diagnostic network optimisation (DNO) analysis using OptiDx was conducted across eight districts in Sumba, North Sumatra, and Central Java, representing high-, moderate-, and low-transmission settings. The maximum annual number of malaria cases from surveillance data between 2019 and 2022 was used as a proxy for testing demand. In baseline scenario, all public health facilities were equipped with G6PD testing devices. In referral scenario, devices were allocated only to selected facilities, with a maximum sample transport distance of 15 kilometres and sample delivery scheduled three times per week. Model outputs were reviewed with the National Malaria Control Programme (NMCP) to identify the optimal scenario, considering factors including implementation cost and accessibility.

**Results:** Equipping 39% (30/76) of facilities in Sumba with G6PD testing devices could reduce total costs by nearly 30% in the first year and approximately 20% in subsequent years compared with the baseline, although the average referral distance increased from 4.48 kilometres to 9.8 kilometres. In North Sumatra, deploying 15 of 68 devices (19%) could reduce total costs by almost 60% in the first year and approximately 15% thereafter. Two health facilities (Puskesmas Padewatu in Sumba and Abdul Manan Hospital in North Sumatra) could experience workload increase exceeding 400 hours annually as referral facilities. In Purworejo, a single device placed at Puskesmas Mranti could enable on-site diagnosis for 56% of patients.

**Interpretation:** The findings demonstrated that DNO can identify cost-efficient and equitable G6PD testing strategies to support safe radical cure and accelerate malaria elimination.

## Background

The Asia-Pacific region has committed to eliminating malaria by 2030. A major barrier to this goal is the relapsing behaviour of *P. vivax*, with the majority of clinical infections in elimination areas being caused by these dormant liver parasites called hypnozoites^1^. Patients can suffer repeated clinical episodes, which lead to cumulative morbidity and also undermine efforts to eliminate transmission. Standard radical cure regimen with 8-aminoquinoline drugs administered over 7 to 14 days kills the hypnozoites and prevents these relapses^2^, but these treatments are contraindicated for patients with glucose-6-phosphate dehydrogenase (G6PD) deficiency. This enzyme deficiency can lead to drug-induced haemolysis, which ranges in severity from being self-limiting to requiring blood transfusion^3^. The World Health Organization (WHO), therefore, recommends routine G6PD testing prior to administering *P. vivax* radical cure^4^, yet access to such testing remains limited, including in Indonesia.

In the absence of routine G6PD testing, health providers are faced with a trade-off between relapse risk and haemolytic risk. Studies have shown that high dose, short course primaquine (7 mg/kg body weight) is the most effective radical cure, whereas in most endemic regions lacking G6PD testing, patients are given low dose 14 days regimen (3.5 mg/kg body weight) to minimise the risk of haemolysis^5,6^. Although relatively safe for individuals with G6PD deficiency, the regimens with lower total doses given over a longer duration are associated with poor adherence^7,8^ and lower efficacy^9^. Females with intermediate G6PD deficiency also have a higher risk of haemolysis with higher daily doses of radical cure primaquine^10^. The introduction of G6PD diagnostics would enable broader use of more effective treatment regimens, reduce relapse rates, and accelerate *P. vivax* elimination. This need is further reinforced by the development of tafenoquine, a single-dose 8-aminoquinoline that requires quantitative G6PD testing prior to administration^11^. Widespread access to G6PD testing would therefore expand access to effective radical cure, reduce the burden of relapse on patients, and limit onward transmission.

Indonesia is one of the malaria hotspots in the Asia-Pacific region, with an estimated 1.1 million cases and 1.9 thousand deaths in 2023 according to the WHO^12^. The country has also committed to ambitious elimination of malaria by 2030. Between 2010 and 2021, Indonesia achieved a 35% reduction in malaria cases; however, relapsing malaria remains a significant challenge, with marked spatial heterogeneity across districts^13^. *P. vivax* mono-and mixed infections account for the majority of relapsing malaria cases, particularly in low- and moderate-endemicity areas. Despite this, routine G6PD screening in patients with acute *P. vivax* or *P. ovale* malaria is typically unavailable, largely due to the need for specialised laboratory skills and equipment and the high cost test to cover a lot of cases, especially in rural settings^14^. In response, the National Malaria Control Programme (NMCP) has prioritised introducing additional interventions to strengthen radical cure strategies and accelerate elimination.

To advance these efforts, the strategic placement and availability of G6PD testing devices are essential, particularly in areas with a high burden of *P. vivax* malaria and a substantial prevalence of G6PD deficiency. Diagnostic network optimisation (DNO) approach, a geospatial network analytics method, can be used to assess diagnostic capacity and recommend the optimal number, placement, and referral pathways for testing services^15^. One of DNO tools is OptiDx, which has been utilised in Kenya and Nepal to design bacteriology and antimicrobial resistance surveillance networks^16^. This study aimed to support the Indonesian NMCP by providing an evidence-based framework to guide the introduction of G6PD diagnostics, optimise resource use, and inform procurement planning and budget advocacy, both domestically and with external funding partners using OptiDx.

## Methods

OptiDx is a web-based DNO analytical tool developed by FIND (Foundation for Global Alliance for Diagnostics)^17^. OptiDx applies a cost-minimisation approach to optimise network design while adhering to predefined constraints^18^. The constraints considered in this study were service distance, which defines the maximum distance a sample can be sent for testing, and the cost to run the testing. OptiDx analysis involved several steps: defining the scope, entering data into the model, building the baseline scenario, evaluating alternative scenarios, and assessing model outputs. Key aspects of this study, such as location selection, model assumptions, and restrictions, were developed collaboratively with NCMP.

### Study settings and scope

The scope of this study was to establish a G6PD testing network. Eight districts from East Nusa Tenggara, North Sumatra, and Central Java provinces were included to represent the range of malaria epidemiological settings^19^ and G6PD prevalence (Figure 1). The malaria high-endemicity setting was represented by four districts in East Nusa Tenggara province (Southwest Sumba, West Sumba, Central Sumba, East Sumba) on Sumba Island, with a combined population of 779,049 in 2020. The prevalence of G6PD deficiency and its mutations in Sumba coastal populations has been well studied, with prevalence ranging from 3.1 to 6.7%, and the most common variants being the severe G6PD Vanua Lava and Viangchan mutations^20^.

**Figure 1.**
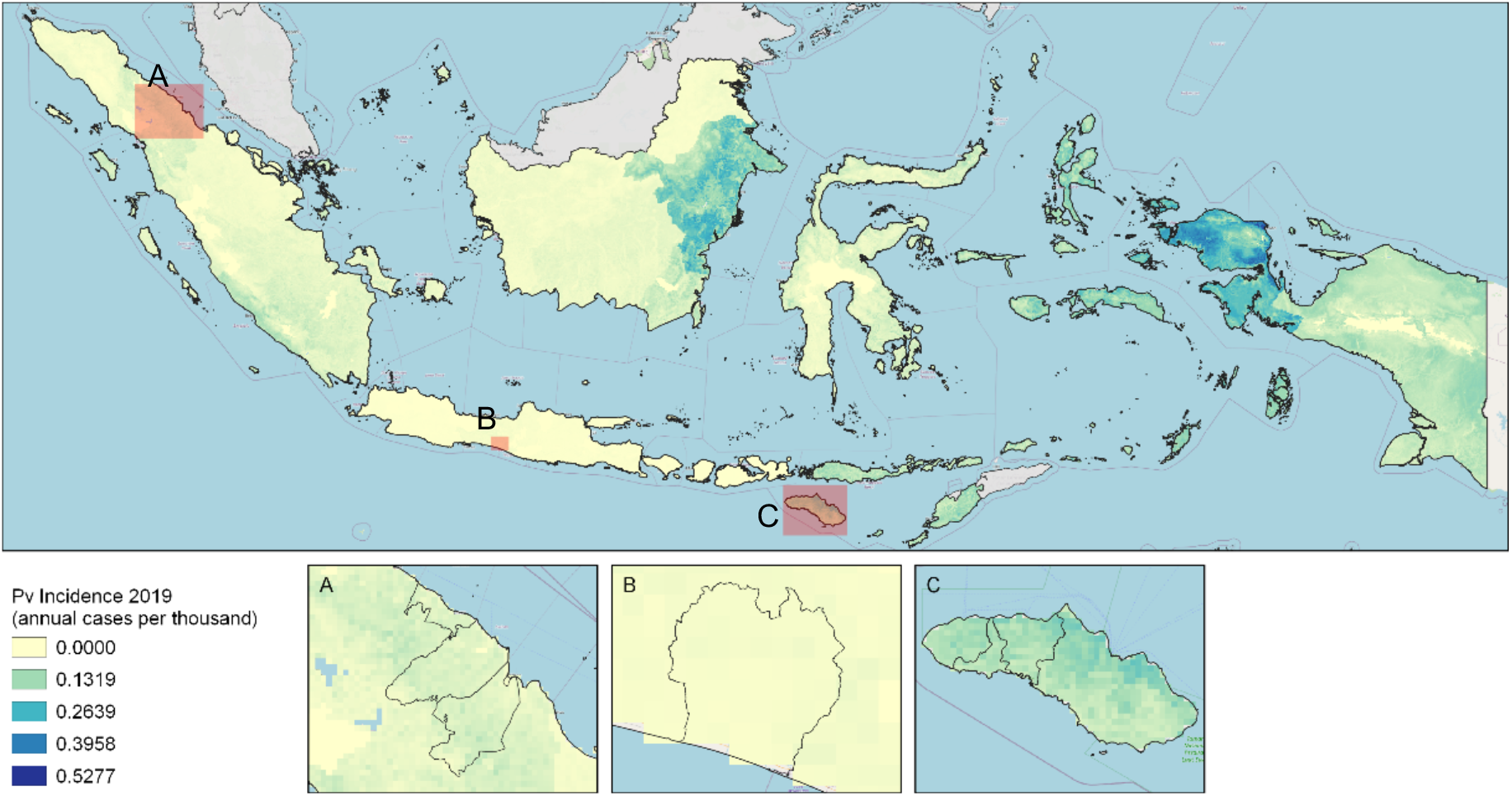
Malaria endemicity in Indonesia based on annual P. vivax cases (2019) in A) North Sumatra Province, *B) Purworejo District, C) Sumba Island. Source: Battle et al. Lancet. 2019*^19^.

Three districts in North Sumatra province, which were Asahan, North Labuhan Batu, and Batu Bara, were selected to represent low-to-moderate transmission settings, with a total population of 1,562,632 in 2020. Purworejo district in Central Java represented a very low transmission, with 2–62 cases reported annually from 2019-2022 and a total population of 769,880 in 2020. Upon review of the reported cases distribution in Purworejo (Figure 2), the relapsing malaria cases were clustered in a similar area every year. Given the limited spatial extent of G6PD testing demand, the data were not considered suitable for a full analysis in OptiDx. However, the cost estimation for installing the device in the health facilities was presented in the results section.

**Figure 2.**
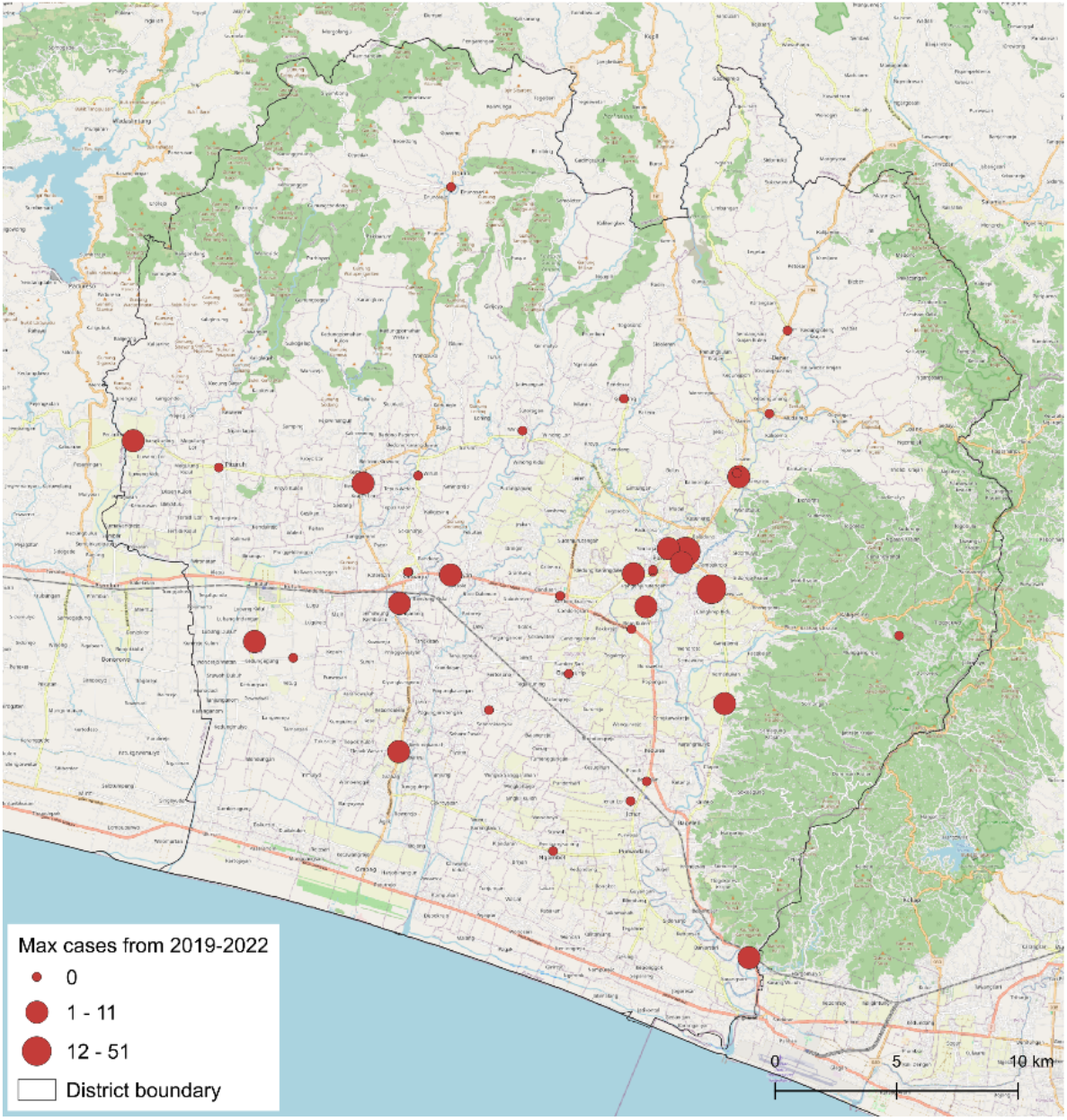
Distribution of the highest reported relapsing malaria cases from 2019 to 2022 by health facility in Purworejo District, Central Java. Baseline map data from OpenStreetMap.

There were three types of health facilities reporting to the malaria surveillance system, including Puskesmas (the primary healthcare [PHC], each subdistrict has at least one), private clinics, and hospitals. In this study, only public health facilities, such as Puskesmas and public hospitals, were considered as potential G6PD testing sites. GPS coordinates for health facilities were searched online using Google and Google Maps. One private clinic in Sumba (BP Bunda Nirmala), which reported up to 246 cases, was excluded due to the unavailability of GPS data. In total, 76 health facilities in Sumba, 68 in North Sumatra, and 38 in Purworejo were included in this analysis.

### Analysis scenarios and assumptions

There were two scenarios analysed in this study: baseline and referral. In both scenarios, samples from patients seeking care at private clinics must always be delivered to the nearest public health facility with a G6PD device. First, the baseline scenario assumed that every public facility was equipped with a G6PD testing device. In the referral scenario, G6PD devices assumed to be allocated only to selected public health facilities, with a maximum one-way referral distance of 15 kilometres, approximately a 30-minute drive by motorised bike. We accounted for the round-trip for each transport, which was a maximum of 30 kilometres and an hour per trip. It was also assumed that healthcare staff at facilities without a G6PD device would need to pool multiple samples up to a maximum of three days after blood sampling. This means that the samples would be delivered approximately three times per week (calculated as 7 days ÷ 3 days per delivery, rounded up) to the nearest public health facility with a G6PD testing device.

### Data collection and inputs

#### Test demand

The reported number of relapsing malaria cases, which were *P. vivax* (mono- and mixed infections) and *P. ovale* cases, at each health facility corresponds to proxy for the G6PD testing demand. Routine malaria surveillance data were obtained from the SISMAL (Malaria Surveillance Information System) NMCP database from 2019 to 2022. The yearly number of cases in each district is shown in Table 1.

**Table 1.** Total reported cases of relapsing malaria (P. vivax mono- and mixed-infection, and P.ovale mono-infection) from 2019-2022 for each province.

| **Province** | **District** | ***P. vivax* mono-infections** |  |  |  | **Mixed infections** |  |  |  | ***P. ovale* mono-infections** |  |  |  |
|---|---|---|---|---|---|---|---|---|---|---|---|---|---|
|  |  | **2019** | **2020** | **2021** | **2022** | **2019** | **2020** | **2021** | **2022** | **2019** | **2020** | **2021** | **2022** |
| **East Nusa Tenggara** | Sumba Barat Daya | 919 | 2260 | 1597 | 2181 | 123 | 318 | 604 | 808 | 0 | 5 | 0 | 1 |
|  | Sumba Barat | 790 | 1342 | 834 | 719 | 19 | 122 | 72 | 205 | 0 | 0 | 0 | 1 |
|  | Sumba Tengah | 58 | 50 | 14 | 14 | 2 | 0 | 1 | 2 | 0 | 0 | 0 | 0 |
|  | Sumba Timur | 226 | 155 | 137 | 429 | 251 | 238 | 182 | 721 | 0 | 0 | 0 | 0 |
| **North Sumatra** | Asahan | 97 | 129 | 784 | 2066 | 0 | 0 | 0 | 4 | 0 | 0 | 0 | 0 |
|  | Labuan Batu Utara | 251 | 217 | 434 | 717 | 0 | 0 | 0 | 0 | 0 | 1 | 0 | 0 |
|  | Batu Bara | 343 | 347 | 700 | 421 | 110 | 102 | 136 | 163 | 0 | 0 | 0 | 0 |
| **Central Java** | Purworejo | 40 | 4 | 1 | 61 | 0 | 0 | 1 | 1 | 0 | 0 | 0 | 0 |

Given that malaria transmission is known to fluctuate in absolute numbers between years and sites, an upper bound for the expected number of relapsing malaria cases was used for more conservative estimates. This was defined as the maximum number of reported annual cases per health facility over 2019–2022, allowing for potential underdiagnosis or underreporting by health facilities.

#### Costs

The OptiDx analysis calculated the costs associated with each diagnostic network scenario. Cost model input data were compiled in consultation with the NMCP team and sourced from national cost references, including the Ministry of Health E-Katalog version 5.0^21^ and the Ministry of Finance budgetary guidance^22^. All costs were searched in Indonesian Rupiah (Rp) as of June 2023, without any forecasting. The average exchange rate in June 2023 was Rp15,007 per US Dollar ($)^23^. Material and training costs were varied in each region, and costs were categorised as follows:

- Test cost: This includes the supplies and consumables required to run a test. This cost will depend on the number of tests done.
- Device operational cost: This includes the device fixed cost, such as the cost of equipment, quality assurance, and refresher training that is set to take place every 6 months.
- Start-up cost: This includes the cost for setting up the device for the first time and initial training for the healthcare facility staff. This cost will not be accounted for in subsequent years.
- Transportation cost: This calculates the fuel per kilometre using a motorised bike, which is the most frequent mode of transport used in the region.

Human resources (HR) were not directly accounted for in the G6PD testing costs, as staff salaries were already paid, and there would be no additional HR charge to the NMCP budget for implementing the G6PD test. However, given the crucial role of health facility staff, including analysts, technicians, clinicians, and nurses, for implementing this network, estimates of the time spent by health workers using the assay were evaluated instead. Each test was estimated to take an average of 15 mins of staff time. The costing details input for analysis were available in Table 2.

**Table 2.** Costing inputs entered into OptiDx.

| Item | Site | Unit cost (IDR) | Pack size | n/ test | Working years | Notes | Source |
|---|---|---|---|---|---|---|---|
| **Test cost: cost per test** |  |  |  |  |  |  |  |
| Standard G6PD test kit | Sumba | 2,473,930 | 25 | 1 | - | Inc. VAT and shipping; inc. 10% failure rate | E-Katalog (June 2023) |
|  | Sumatra | 2,378,930 |  |  | - |  |  |
| Nitrile gloves |  | 125,000 | 100 | 2 | - | Inc. shipping cost |  |
| Alcohol swab |  | 37,754 | 100 | 1 | - | Inc. shipping cost |  |
| Lancet |  | 40,000 | 200 | 1 | - | Inc. shipping cost |  |
| EDTA tubes 0.4 ml |  | 58,100 | 50 | 1 | - | Inc. shipping cost |  |
| Syringe 1 cc |  | 124,168 | 100 | 1 | - | Inc. shipping cost |  |
| **Device operational cost** |  |  |  |  |  |  |  |
| **Overhead cost: Equipment** |  |  |  |  |  |  |  |
| G6PD Analyser (back-up) | Sumba | 7,183,800 | - | - | 3 | Inc. VAT and shipping | E-Katalog (June 2023) |
|  | Sumatra | 7,088,800 | - | - |  |  |  |
| Standard G6PD control test | Sumba | 830,200 | - | - | 0.2 | Inc. VAT and shipping |  |
|  | Sumatra | 735,200 | - | - |  |  |  |
| Battery for device (AAA) |  | 51,000 | - | - | 0.25 | Inc. shipping<br>Changed every 3 months |  |
| Lancing device |  | 17,914 | - | - | 10 | Inc. shipping |  |
| Cool box | Sumba | 2,701,200 | - | - | 10 | Inc. shipping |  |
|  | Sumatra | 2,478,900 | - | - | 10 | Inc. shipping |  |
| Ice Pack |  | 17,000 | - | - | 3 | Inc. shipping |  |
| **Overhead cost: Quality** |  |  |  |  |  |  |  |
| Annual refresher training | Sumba | 10,112,060 | - | - | - |  | SBU<sup>†</sup> 2023 |
|  | Sumatra | 8,615,560 | - | - | - |  | SBU<sup>†</sup> 2023 |
| **Start-up cost** |  |  |  |  |  |  |  |
| Standard G6PD test kit | Sumba | 2,261,300 | - | - | 1 | Inc. VAT and shipping | E-Katalog (June 2023) |
|  | Sumatra | 2,166,300 | - | - |  |  |  |
| G6PD Analyser | Sumba | 7,183,800 | - | - | 3 | Inc. VAT and shipping |  |
|  | Sumatra | 7,088,800 | - | - |  |  |  |
| Initial training | Sumba | 6,463,530 | - | - | 1 |  | SBU<sup>†</sup> 2023 |
|  | Sumatra | 5,638,180 | - | - |  |  |  |
| **Transportation cost (health system-incurred cost)** |  |  |  |  |  |  |  |
| Transport (motorbike) |  | 20,000 | - | - | - | Per km (to and from) | NMCP meeting |

<sup>†</sup> Ministry of Finance budgetary guidance (General Cost Budget [*Standar Biaya Umum*/SBU])

### Distances between healthcare facilities and transport accessibility

OptiDx measures the referral distances using the straight-line distance method, adjusting for a network correction factor of 40%^24^. This method does not account for the impact of travel barriers such as mountains and water bodies, or the presence of straight roads, which may reduce actual travel distances relative to the OptiDx estimate. To address these limitations, more realistic accessibility analysis for the sample referral travel network was conducted using AccessMod, which incorporates road network coverage, landcover types, digital elevation model (DEM), water bodies, and other barriers^25^. Travel speeds were estimated based on the DEM and landcover types, with walking and motorbike travel speeds referenced from Weiss et al.^26^ and Liecht et al.^27^. Details on the analysis using AccessMod were provided in the Additional file 1.

### Selection criteria for DNO model outputs

The model outputs were evaluated together with NMCP to determine the optimal scenario. Several factors were considered in the final model, including the proportion of health facilities with on-site devices, the accessibility of the nearest health facilities with the testing device, the cost per test, and the total implementation cost. Additionally, the annual number of diagnostic tests and network efficiency were assessed. Network efficiency is evaluated based on the cost outcome, which is calculated as the total diagnostic network cost divided by the number of tests performed, and utilisation, measured as the number of tests performed divided by the maximum device testing capacity. Furthermore, equitable distribution of geographic access among populations, particularly in hard-to-reach communities, were considered.

## Results

### Sumba

The model data included 7179 relapsing malaria cases for analysis. In the baseline scenario, devices were allocated to 89% (68/76) of health facilities reporting relapsing malaria cases on Sumba Island. The remaining 11% (8/76) were private clinics that would need to refer blood samples to the nearest public health facility with G6PD testing available. The total cost to the NMCP in the first year was estimated at Rp2,765,885,000 ($184,306), of which 27% was allocated to system set-up. This corresponded to an average cost of Rp385,000 ($26) per *P. vivax* case. In the subsequent years, the cost could decrease by 27%, resulting in an annual cost of Rp2,009,800,000 ($133,924) and an average cost of Rp280,000 ($19) per case.

On the other hand, the referral scenario allocated only 39% G6PD devices (30/76), which substantially reduced start-up costs by about 56% but increased transportation costs by fourfold. The total cost to the NMCP in the first year was estimated at Rp1,985,308,000 ($132,292), with an average cost of Rp277,000 ($18) per case. We anticipated a 17% reduction in subsequent years, with a total annual cost of Rp1,651,741,000 ($110,065) and an average cost of Rp230,000 ($15) per case. The largest cost component in the referral scenario was test cost (38%), while in the baseline scenario, it was device operational cost (42%). Overall, the referral scenario reduced total costs by almost 30% in the first year and approximately 20% in subsequent years compared with the baseline scenario. The detailed summary of each G6PD network scenario is shown in Table 3.

**Table 3.** DNO model output for each G6PD network scenario in Sumba and North Sumatra.

|  | **Sumba** |  | **North Sumatra** |  |
|---|---|---|---|---|
|  | Baseline | Referral<sup>†</sup> | Baseline | Referral<sup>†</sup> |
| Recommended G6PD device number | 68 | 30 | 78 | 15 |
| Total cost to NMCP in the start-up year<sup>§</sup> | 2,765,885 | 1,985,308 | 2,373,076 | 954,927 |
| Total running cost to NMCP in subsequent years<sup>§</sup><br>(excluding start-up cost) | 2,009,800 | 1,651,741 | 1,580,066 | 802,425 |
| Cost components |  |  |  |  |
| Total start-up cost<sup>§</sup> | 756,085 | 333,567 | 793,010 | 152,502 |
| Total test cost<sup>§</sup> | 749,732 | 749,732 | 401,228 | 401,228 |
| Total device op cost<sup>§</sup> | 1,165,407 | 514,150 | 1,178,838 | 226,700 |
| Sample transport cost<sup>§</sup> | 94,661 | 387,859 | 0 | 174,498 |
| Cost per case in the start-up year<sup>§</sup> | 385 | 277 | 595 | 240 |
| Cost per case in subsequent year<sup>§</sup> | 280 | 230 | 396 | 201 |
| Average referral distance | 4.48 km | 9.8 km | 0 km | 2.8 km |
| % sample per travel distance interval<br>(km, two-way): |  |  |  |  |
| 0–2 | 73.0 | 42.2 | 100 | 42.5 |
| 2–10 | 4.7 | 8.9 | 0 | 36.8 |
| 10–20 | 16.3 | 34.4 | 0 | 15.4 |
| 20–30 | 6.0* | 14.5 | 0 | 5.2 |
| Average device utilisation | 1.32% | 2.99% | 0.64% | 3.32% |

<sup>†</sup> Max referral distance: 15km, one-way travel time by motor bike ± 30 min
<sup>§</sup> In thousand IDR. Costs and distances account for return trips.
<sup>\*</sup> The estimated 6% of patient samples had to travel 20-30km (beyond the maximum limit evaluated in this scenario) were from private clinics, where the introduction of G6PD tests is not under consideration.

The average round-trip travel distance for private clinics to deliver samples to the nearest facility equipped with a G6PD device was 4.48 km in the baseline scenario, with most (73%) of the samples requiring to travel 2 kilometres or less. In contrast, under the referral scenario, facilities requiring sample transport had a higher mean travel distance of 9.8 kilometres, with slightly above 50% of cases travelling 2-10 kilometres. Figure 3 illustrates the relationship between sample referral distance and transportation costs, stratified by the number of cases. In general, greater distances between referring and testing facilities led to higher transportation costs. These costs were further influenced by case volume, as facilities with a higher number of cases required more frequent sample referrals, resulting in higher travel expenses. Notably, three health facilities reporting 120–149 cases required round-trip travel distances of 20 kilometres or more.

**Figure 3.**
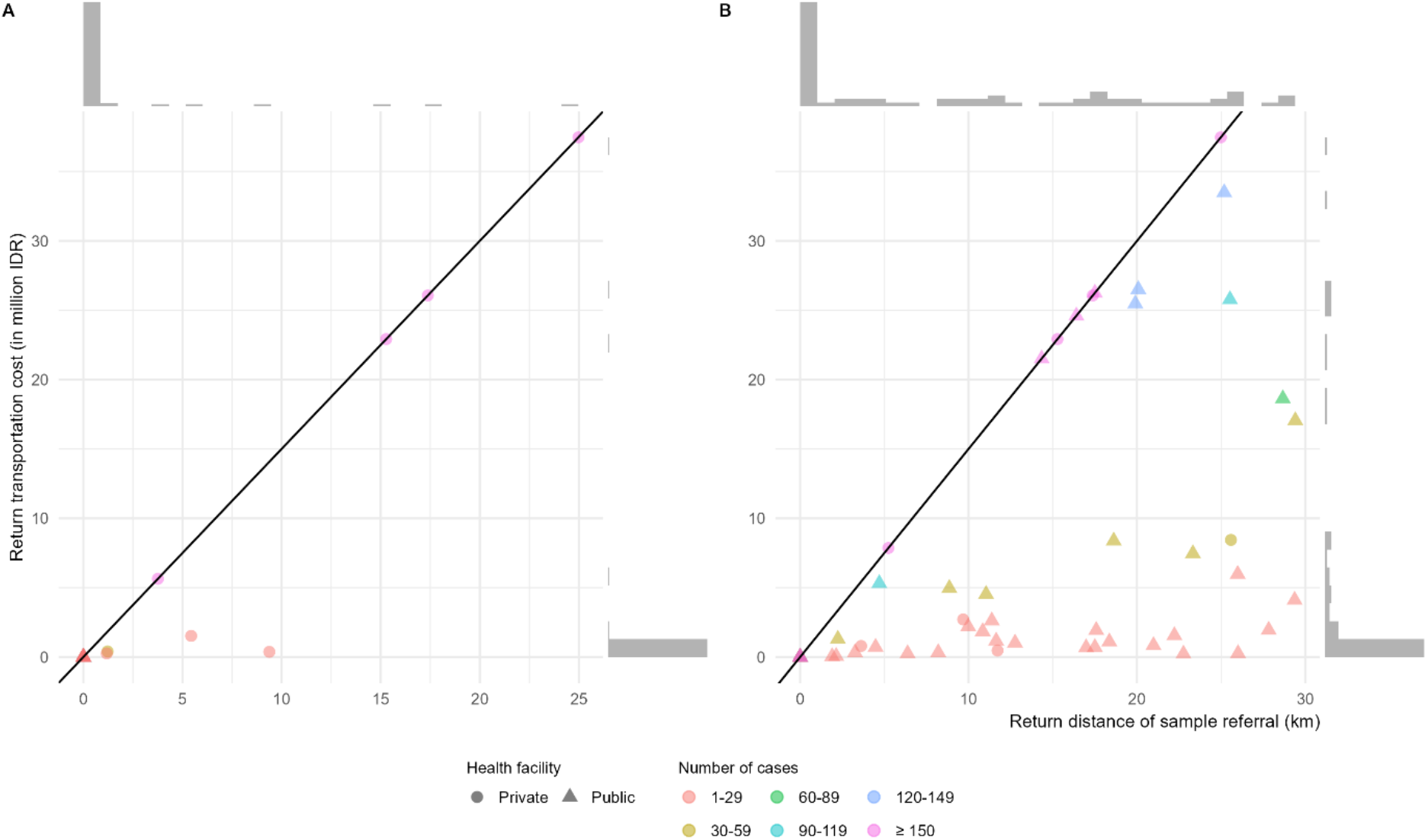
Distance of sample referral in association with transportation cost for all cases for A) baseline and B) referral scenarios on Sumba Island. The diagonal line represents the cost ceiling, where higher costs along this line only correspond to longer distances (not to the number of cases, due to sample pooling). Below this line, transportation costs are determined by both distance and the number of cases.

Figure 4 illustrates the geographical distribution of G6PD devices in the baseline and referral scenarios. The additional workload associated with introducing an extra step in the management of relapsing malaria, whether due to the time required for testing or for travel to transport sample, is shown in Figure 5. Although overall device utilisation was low (1.32% in the baseline scenario and 2.99% in the referral scenario), and no device exceeded its testing capacity, Puskesmas Padewatu was required to accommodate a substantial increase in workload, with an additional 300 and 400 hours of work annually for G6PD testing in the baseline and referral scenarios, respectively.

**Figure 4.**
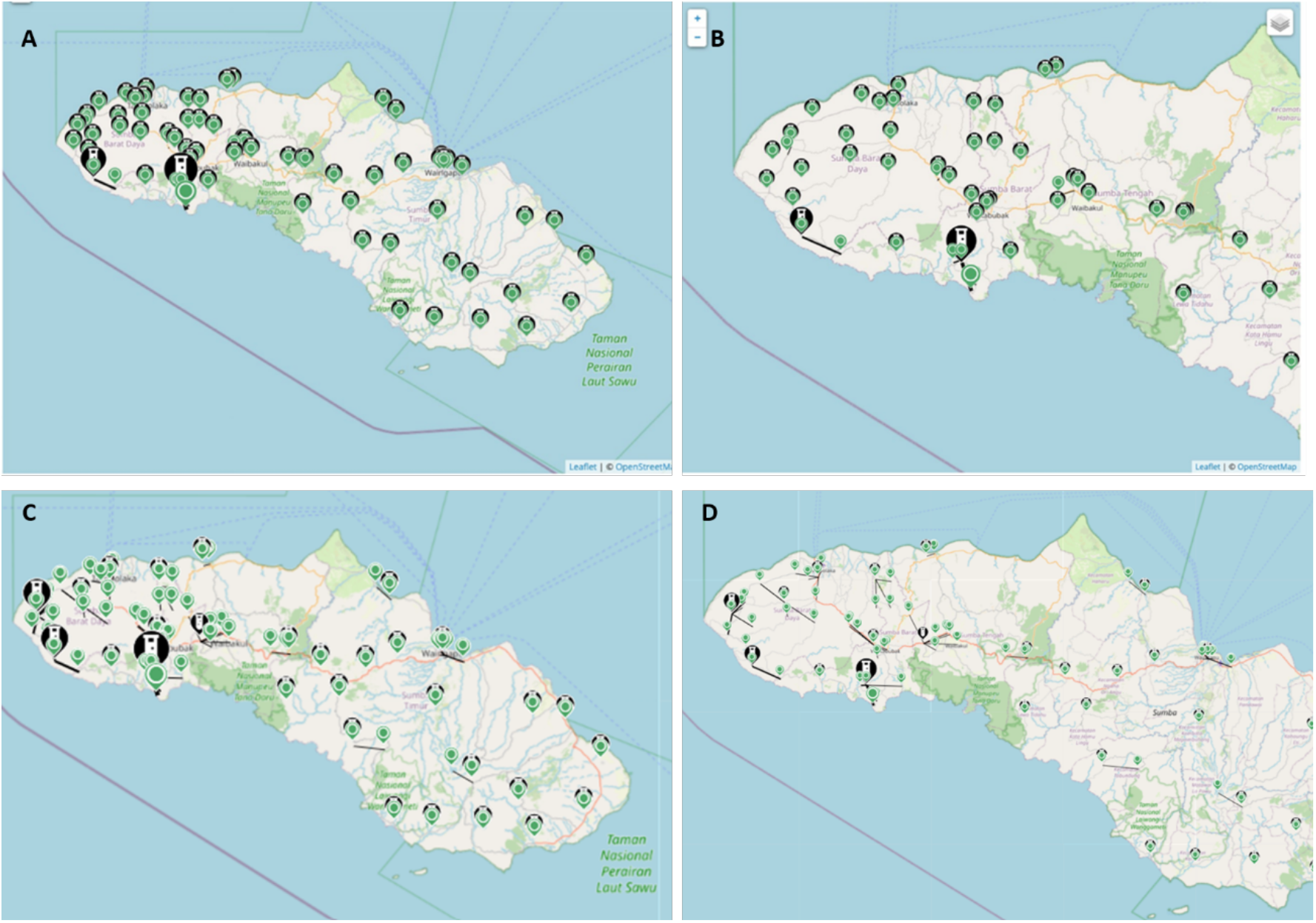
Map showing the allocation of G6PD devices in the baseline scenario on Sumba Island A) overall view, B) zoomed-in view, and in referral scenario on Sumba Island, C) overall view, D) zoomed-in view. Green circles represent health facilities that reported P. vivax cases. Health facilities marked with a tube symbol inside a black circle indicate that they have been allocated a G6PD device. Solid line represents a referral connection between a facility that needs to transport samples and a facility with a device.

**Figure 5.**
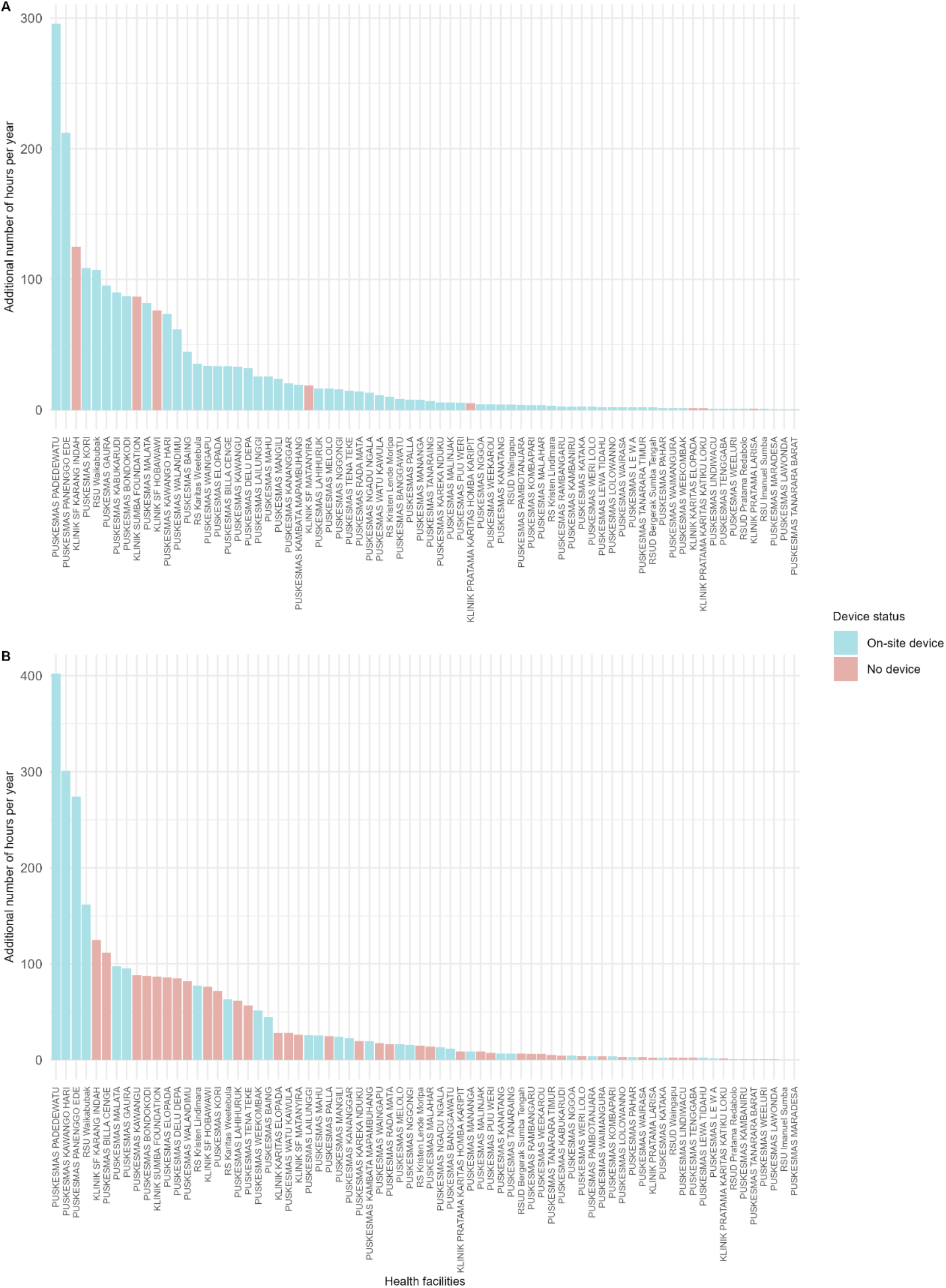
Additional hours per year required for G6PD testing in each health facility on Sumba Island under the A) baseline and B) referral scenarios. Additional time worked at facilities without a device was due to travel, whereas at facilities with an on-site device, it was due to testing.

### North Sumatra

The model data included 3987 relapsing malaria cases for analysis. In the baseline scenario, G6PD devices were allocated to all health facilities (n = 68) reporting relapsing malaria cases in North Sumatra. The total first-year cost to the NMCP was estimated at Rp2,373,076,000 ($158,131), of which 33% was attributable to system setup, corresponding to an average cost of Rp595,000 ($40) per case. In the subsequent years, the cost decreased by 33%, resulting in an annual cost of Rp1,580,066,000 ($105,289) and an average cost of Rp396,000 ($26) per case.

On the other hand, the referral scenario allocated only 19% G6PD devices (15/68), substantially reducing start-up costs by about 81%, but introducing an additional transportation cost of Rp174,498,000 ($11,628). The total cost to the NMCP in the first year was estimated at Rp954,927,000 ($63,632), equivalent to an average cost of Rp240,000 ($16) per case. The cost in subsequent years could decline by 16%, resulting in an annual total cost of Rp802,425,000 ($53,470) or Rp201,000 ($13) per case. The largest cost component in the referral scenario was test cost (50%), while in the baseline scenario, it was device operational cost (50%). Overall, the referral scenario reduced total costs by almost 60% in the first year and approximately 15% in subsequent years compared with the baseline scenario. The detailed summary of each G6PD network scenario is shown in Table 3.

Under the referral scenario, 81% of facilities (63/78) were required to refer blood samples to the nearest public health facility with G6PD testing available. These facilities had an average round-trip travel distance of 2.8 kilometres, with just above 40% travelling 2 kilometres or less. Figure 6 illustrates the relationship between sample referral distance and transportation costs, stratified by the number of cases. Notably, two health facilities reporting 120–149 cases required round-trip travel distances of nearly 20 kilometres.

**Figure 6.**
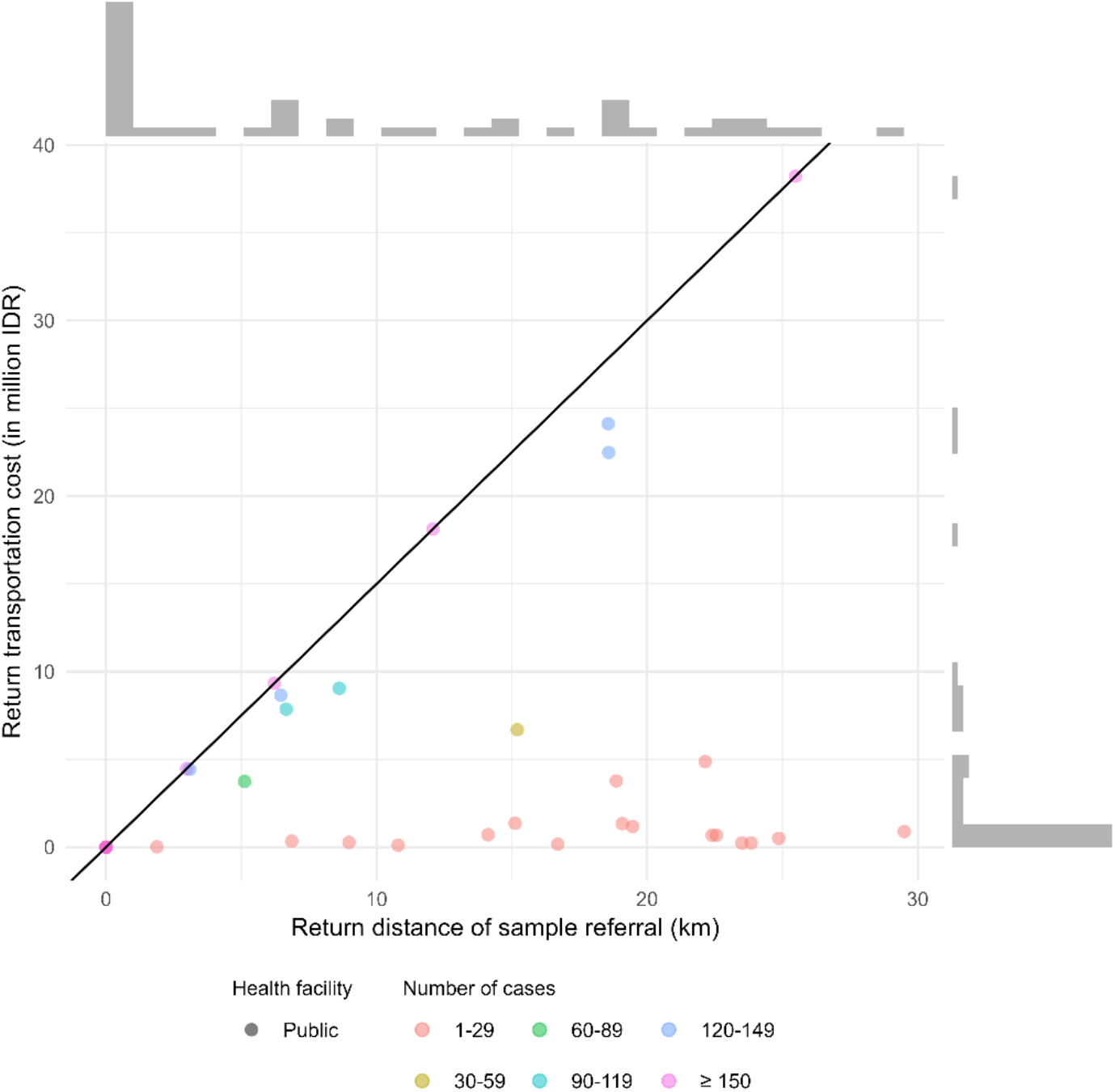
Distance of sample referral in association with transportation cost for all cases for the referral scenario in North Sumatra. The diagonal line represents the cost ceiling, where higher costs along this line only correspond to longer distances (not to the number of cases, due to sample pooling). Below this line, transportation costs are determined by both distance and the number of cases.

Figure 7 illustrates the geographical distribution of G6PD devices in the baseline and optimised referral scenarios. The additional workload associated with introducing an extra step in the management of relapsing malaria, whether due to the time required for testing or for travel to transport sample, is shown in Figure 8. Although overall device utilisation was low (0.64% in the baseline and 3.32% in the referral scenario), and no device exceeded its capacity, Puskesmas Tanjung Leidong was estimated to have an extra working hour of 150, while Abdul Manan Hospital got more than 400 additional working hours per year after becoming a referral facility.

**Figure 7.**
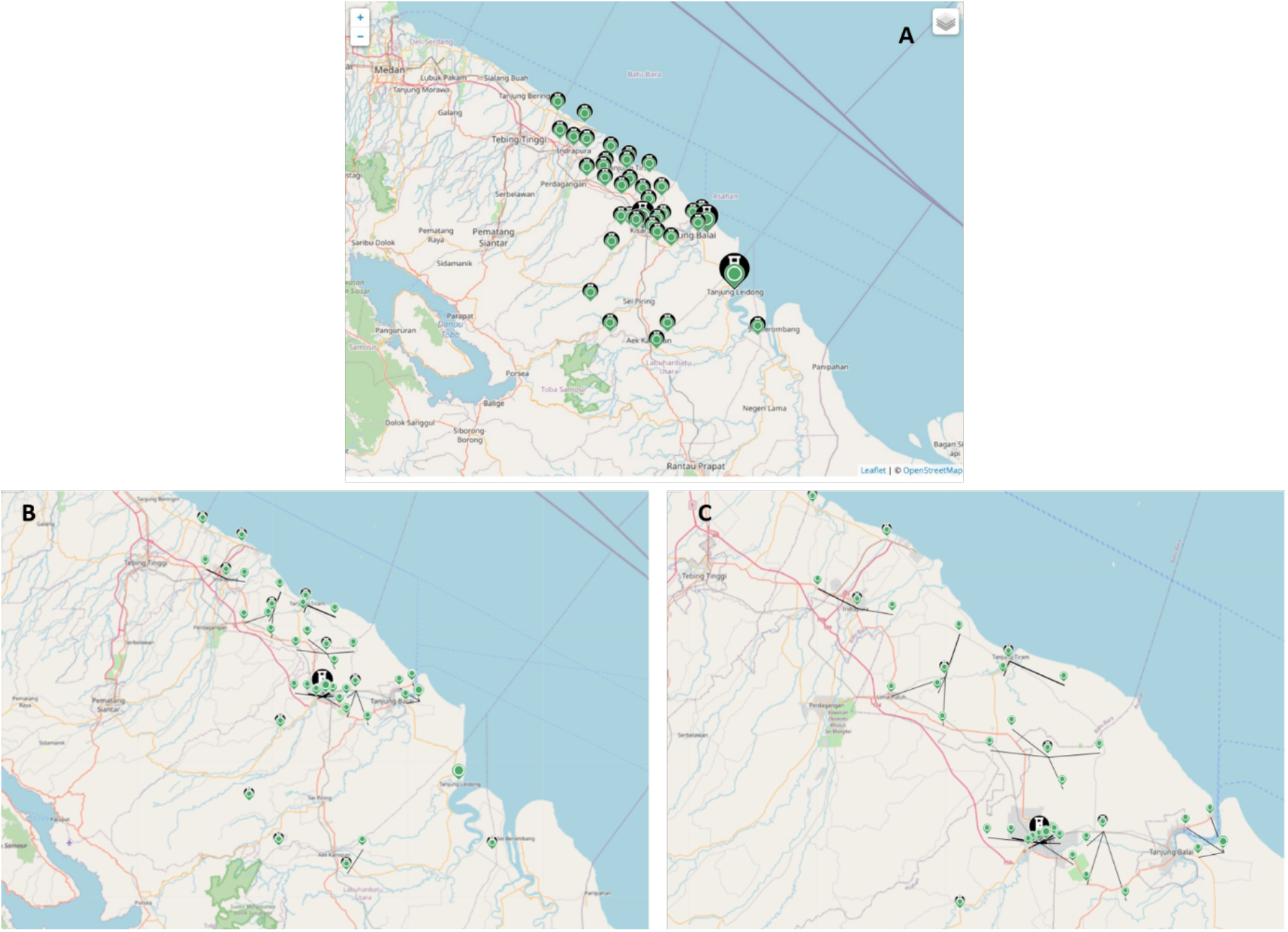
Map showing the allocation of G6PD devices in the A) baseline scenario in North Sumatra, where no transport was required as all facilities were allocated a device, and in the referral scenario in North Sumatra B) overall view, C) zoomed-in view. Green circles represent health facilities that reported P. vivax cases. Health facilities marked with a tube symbol inside a black circle indicate that they have been allocated a G6PD device. Solid line represents a referral connection between a facility that needs to transport samples and a facility with a device.

**Figure 8.**
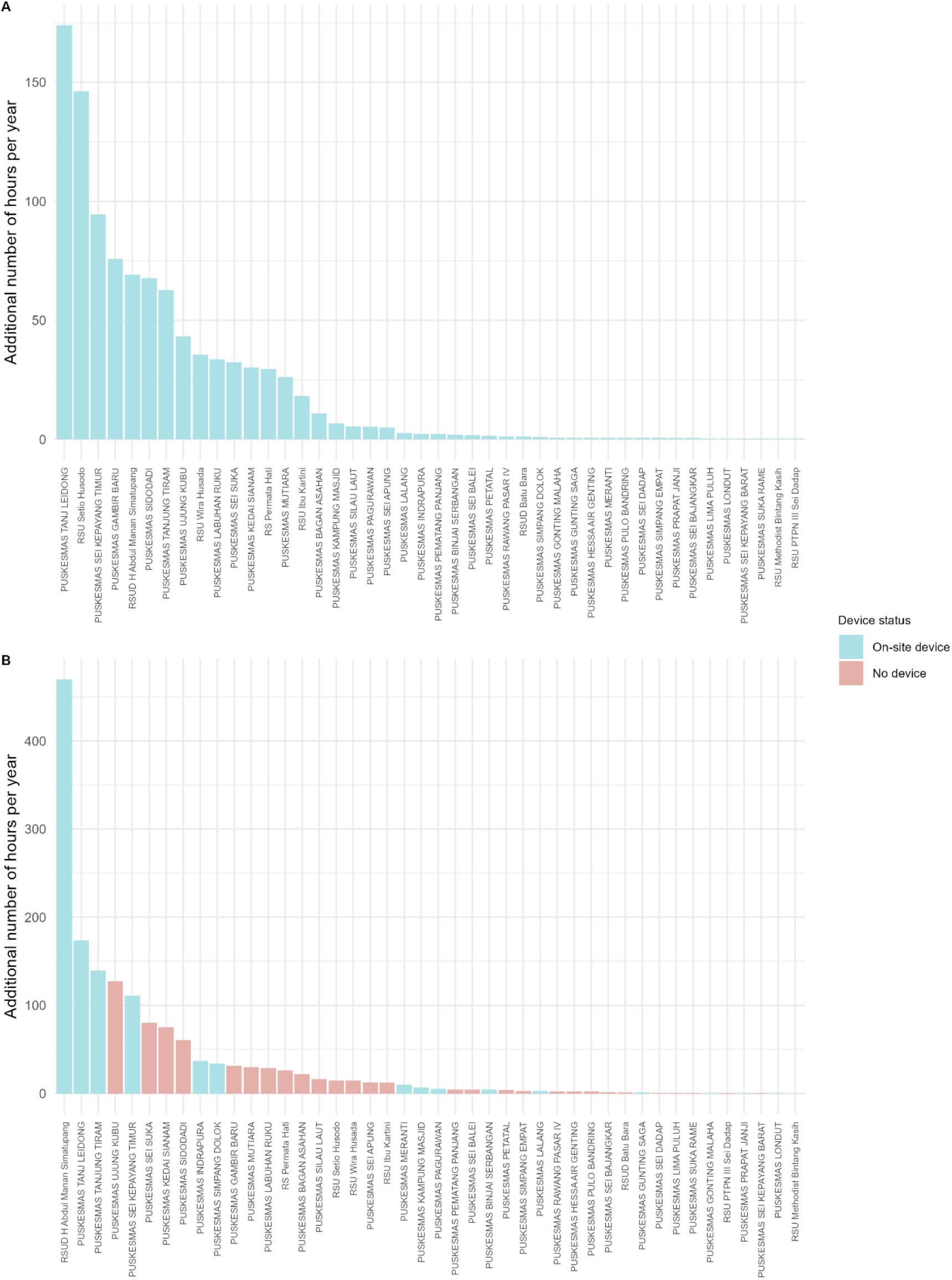
Additional hours per year required for G6PD testing in each health facility in North Sumatra under the A) baseline and B) referral scenarios. Additional time worked at facilities without a device was due to travel, whereas at facilities with an on-site device, it was due to testing.

### Purworejo

Based on the spatial distribution of the cases and an annual caseload of 2–62, introducing G6PD testing capacity at the hotspot health facility, Puskesmas Mranti, would likely be sufficient and would allow around 56% of patients to be treated on-site without referral. The maximum travel distance from Puskesmas Mranti to the most distant reporting health facility with cases (Puskesmas Karangetas site reported 1 case in 2019) was 25.3 km. NMCP could allocate a G6PD testing device to the Puskesmas Mranti health facility at the district centre.

Assuming there were 60 patients per year, the estimated costs would be a start-up cost of Rp14,816,000 ($987), an annual device operational cost of Rp18,671,000 ($1,244), and a test cost of Rp124,000 ($8) per patient. This would total Rp40,892,000 ($2,725) in the first year and Rp26,076,000 ($1,738) in the subsequent years. For the sample referral travel cost, the furthest referral to the central G6PD testing site was approximately 33 km, so the upper limit of the travel cost would be approximately Rp330,000 ($22) per trip.

## Discussion

This study is the first to explore the use of DNO to inform planning for G6PD testing in malaria programme. The results indicate that designating selected health facilities as referral facilities can improve the cost efficiency of G6PD testing implementation while maintaining sufficient population coverage. Including private facilities, especially those with a high incidence of malaria cases, may further increase coverage. In addition to cost, it is important to consider additional workload at health facilities to prevent excessive burdens on healthcare workers. Findings from OptiDx may serve as a pilot empirical framework for future scale-up to broader geographical regions.

While applying DNO in a very low transmission setting with occasional outbreaks, such as in Purworejo, was deemed unreliable, the approach proves highly relevant in moderate to high transmission areas like Sumba Island and North Sumatra. Our analysis suggests that, compared to a baseline scenario, the optimised distance-restricted scenario could substantially reduce NMCP’s expenditure on integrating G6PD testing. In the referral scenario, an estimated 38 and 63 fewer devices were required in Sumba and North Sumatra, respectively, resulting in estimated total cost reductions of 30% and 60% during the start-up year. These efficiencies were achieved while maintaining reasonable sample transport distances for healthcare staff and ensuring that no allocated device exceeds its testing capacity, thereby accommodating potential future increases in test demand. Previous study applyting OptiDx to optimise tuberculosis (TB) diagnostic networks in Kenya, India, and the Phillipines similarly demonstrated its value in supporting evidence-based placement of diagnostic tools. In Kenya, OptiDx was used to optimise utilisation of the Xpert MTB/RIF platform for TB and HIV testing. In India, the model was updated with current programme data to estimate future testing demand and diagnostic capacity. In the Philippines, OptiDx supported the optimal placement of diagnostic devices within the national network^28^.

However, we also observed the significant additional hours placed on healthcare staff as a result of implementing G6PD testing (e.g., time spent on maintaining the device, conducting tests, or transporting samples). This added hours potentially increases the burden on already overstretched staff, particularly in facilities with a high caseload. In selected facilities with a high number of additional hours (e.g., an estimated average of nine or more extra hours per week at Abdul Manan Hospital, North Sumatra), the deployment of additional staffing may be necessary to ensure the sustainability of G6PD testing services without compromising routine healthcare delivery.

While helpful, this analysis and its inherent assumptions are just one component of a more complex, evidence-informed policymaking process. It is a guidance rather than a strict prescription. For example, special considerations for private clinics on southwest Sumba coast could be considered. Three private clinics, which was Klinik Sumba Foundation Karang Indah, Klinik Sumba Foundation, and Klinik Sumba Foundation Hobawawi on the south coast of west Sumba collectively contributed 22% of the total demand for G6PD tests on Sumba. Referral distances from these private clinics to recommended nearby G6PD testing sites were estimated at 12.49 km, 8.69 km, 7.64 km respectively. These patients represented approximately 80% of the patients requiring referral consultations for G6PD testing within the region. Given the uncertain level of referral uptake, it may be considered by the NMCP to position some additional G6PD tests in these facilities, despite them not being public health facilities.

Indonesia has recently considered introducing a new treatment option for *P. vivax* malaria. Under the proposed new guidelines, patients may receive primaquine at a dose of 1 mg/kg body weight daily for 7 days provided G6PD testing returns normal (Indonesian Malaria Diagnosis and Treatment Working Group 2026, unpublished), in addition to 0.25 mg/kg daily for 14 days. This policy change is driven by evidence suggesting that the longer 14-day regimen often results in poor patient adherence, undermining treatment effectiveness and increasing the risk of malaria relapse^8^. By halving the duration of treatment, the new 7-day regimen is expected to improve compliance and treatment completion rates among patients. However, the higher daily dose of primaquine raises significant safety concerns, particularly for individuals with G6PD deficiency.

There were several limitations to this analysis that should be considered before implementation. First, the analysis was limited to evaluating the proximity of health facilities to G6PD testing sites, without accounting for patients’ places of residence, and may have underestimated the costs patients incur to access healthcare. Individuals who do not seek care due to financial or logistical constrains would not be adequately represented in routine surveillance data. In addition, there was underreporting issue in the use of surveillance data, which may have led to an underestimate of testing demand. To mitigate this, the analysis used the maximum site-specific number of *P. vivax* cases rather than average caseloads.

Cost estimates were based on 2023 data and did not account for inflation or future budgetary adjustments, allowing for a relative budget assessment. This analysis also did not consider existing referral systems, such as the Universal Health Coverage by the Healthcare and Social Security Agency tiered referral system, which may influence patient pathways for G6PD testing and/or radical cure treatment. Our analysis also assumed that all health facilities under consideration have the same quality and infrastructure. In reality, facility readiness and resource availability vary, tailored implementation strategies are likely needed for optimal outcomes.

Travel speeds vary and, in this analysis, did not include empirical travel time data specific to the study regions. We used generic travel speed data which may not actually represent the terrain conditions in Sumba Island and North Sumatra. The current analysis is only able to analyse three regions with relatively good road network (i.e., available transport accessibility), omitting areas with challenging terrain where access to healthcare is limited. In the referral scenario, patients would not receive radical cure immediately, creating additional workload for healthcare staff to either notify patients when treatment becomes available or arrage drug delivery. Finally, scalability was constrained by the use of administrative boundaries as analytical units. While regions such as Sumba Island are relatively isolated, demand in North Sumatra is directly influenced by its neighbouring provinces, which are not accounted for in the analysis.

## Conclusion

This study demonstrated that the quantitative optimisation method using DNO is a powerful tool to assist the NMCP in allocating G6PD devices more cost-efficiently while ensuring equity and access to improved healthcare in managing relapsing malaria. Systematically integrating G6PD testing before administering primaquine, and potentially tafenoquine in the future, could reassure patients and clinicians that the benefits of treatment likely outweigh the harms. This helps to improve patient compliance and, consequently, treatment completion. This represents a crucial incremental step in reducing ongoing transmission within the population and, ultimately, achieving elimination. Furthermore, stronger collaboration among researchers, policymakers, and healthcare providers is essential to ensure that optimisation strategies are effectively integrated into health systems. Future analyses may include other high-burden regions, such as Papua, to accelerate case reduction. Additionally, evaluating the costs and benefits of implementing G6PD testing as part of a broader *P. vivax* radical cure programme, using a transmission model, would be valuable for understanding the long-term public health and economic impacts of these interventions.

## Data Availability

The malaria data that support the findings of this study are from the Malaria Working Group, Ministry of Health of Indonesia, but restrictions apply to the availability of these data, which were used under license for this study, and so are not publicly available. Data are however available upon reasonable request and with written approval from the Malaria Working Group, Ministry of Health of Indonesia.

## Acknowledgement

Exeins Health Initiative and OUCRU Indonesia have received a small grant from FIND to conduct this study in Indonesia. FIND is supported by core funding from the Australia Department of Foreign Affairs and Trade (DFAT) and the Swiss Agency for Development and Cooperation (SDC). Funding for OptiDx comes from multiple sources including Bill and Melinda Gates Foundation (BMGF), The U.S. President’s Emergency Plan for AIDS Relief (PEPFAR), USAID and The Global Fund. We are thankful for the technical support provided by Amit Singh, Juhi Gautam and Heidi Albert from FIND.

## Authors’ contributions

IRFE, AWS, RH conceptualised the study. KDL, IF, IN was responsible for coordinating between NMCP and FIND, collating and cleaning datasets, formal analysis, visualisation, and writing the initial draft. HDP, MT, BK, HS, RKH, DP, AKN, MBHP, RTPD, HH provided the scenario selection and model assumptions/criteria, access to required datasets and costings, and reviewed the preliminary outputs to help refine the final analyses. All authors had critically reviewed the study for intellectual content and had full responsibility for the decision to submit the final manuscript.

## Ethics

No ethical committee review was required since no identifiable patient data was accessed and no clinical subjects were involved in the study.

