## Additional file 1 for "Applying Diagnostic Network Optimisation analysis to inform the introduction of G6PD testing into Indonesia for improved treatment of relapsing malaria"

**Distances between healthcare facilities and transport accessibility**

OptiDx measures the referral distances using the straight-line distance method, adjusting for a network correction factor of 40%[1]. This method does not account for the impact of travel barriers such as mountains and water bodies, or the presence of straight roads, which may reduce actual travel distances relative to the OptiDx estimate. To address these limitations, accessibility analysis for the sample referral travel network was conducted using AcessMod 5 (v5.8.0)[2,3], which incorporates road network coverage, landcover types, digital elevation model (DEM), water bodies, and other barriers[2].

Travel time between healthcare facilities was calculated using 'Accessibility' module. We estimated healthcare accessibility at 1 km × 1 km resolution with no restriction of travel time towards healthcare facilities and anisotropic rule. All spatial layers projected into WGS 84 EPSG:3857 projection to have meters as map units. We also measured the referral network which include travel time and distance between the healthcare facilities using 'Referral' module. We estimated referral network at 1 km × 1 km resolution and anisotropic rule. The data we used for this analysis were the road network, water bodies, landcover, and Digital Elevation Model (DEM) downloaded on June 2023 from the Geospatial Information Agency website[4] which was a map repository under Ministry of National Development Planning. Healthcare facilities coordinates was searched online using Google and Google Maps. Travel speeds were estimated based on the landcover types, with reference to the walking and motorbike travel speeds listed in Table Figure S3.

*Table S1 Reference travel speeds according to landcover for input into AccessMod*

| | **Input speed (km/hr)**[†] | | **Equivalent category as estimated in Weiss et al *Nature* (2018)**[5] |
|---|---|---|---|
| Pier | Walking | 5 | Urban |
| Building | Walking | 5 | Urban |
| Forest | Walking | 3 | Mixed forest |
| Savanna | Walking | 5 | Savanna |
| Plantation | Walking | 3 | Average of "cropland" and "cropland vegetation" |
| Settlement | Walking | 5 | Urban |
| Rice field | Walking | 3 | Average of "cropland" and "cropland vegetation" |
| Rainfed agriculture | Walking | 3 | Average of "cropland" and "cropland vegetation" |

| | | | |
|---|---|---|---|
| Shrubland | Walking | 3 | Barren vegetation |
| Dryland agriculture | Walking | 3 | Average of “cropland” and “cropland vegetation” |
| Non-agricultural vegetation | Walking | 3 | Barren vegetation |
| Mangrove | Walking | 2 | Permanent wetlands |
| Sand | Walking | 5 | No equivalent in Weiss et al. 2018<br>Value from Liecht et al. 2007[6] |
| Toll road | Motorbike | 50 | Motorbike road travel speed was estimated from Google Maps travel estimates for each road type |
| Main road | Motorbike | 40 | |
| Secondary road | Motorbike | 40 | |
| Tertiary road | Motorbike | 20 | Road type is not featured in Google Maps, speed estimated to half the main/secondary road speed; speed likely low given poorer road quality |

† Speed rounded to nearest integer

The result from AccessMod were modified in QGIS v3.16.6. Figure S1 and Figure S2 showed the accessibility surface from AccessMod on Sumba and North Sumatra, respectively.

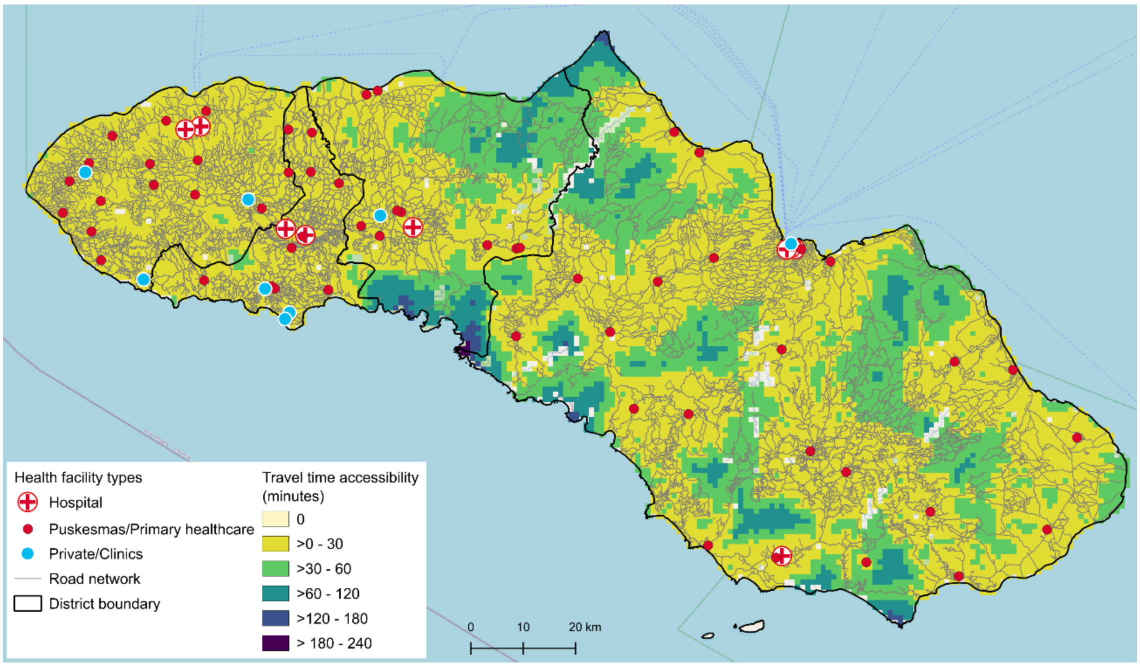

*Figure S1 Accessibility surface from AccessMod with overlaid road network on Sumba Island*

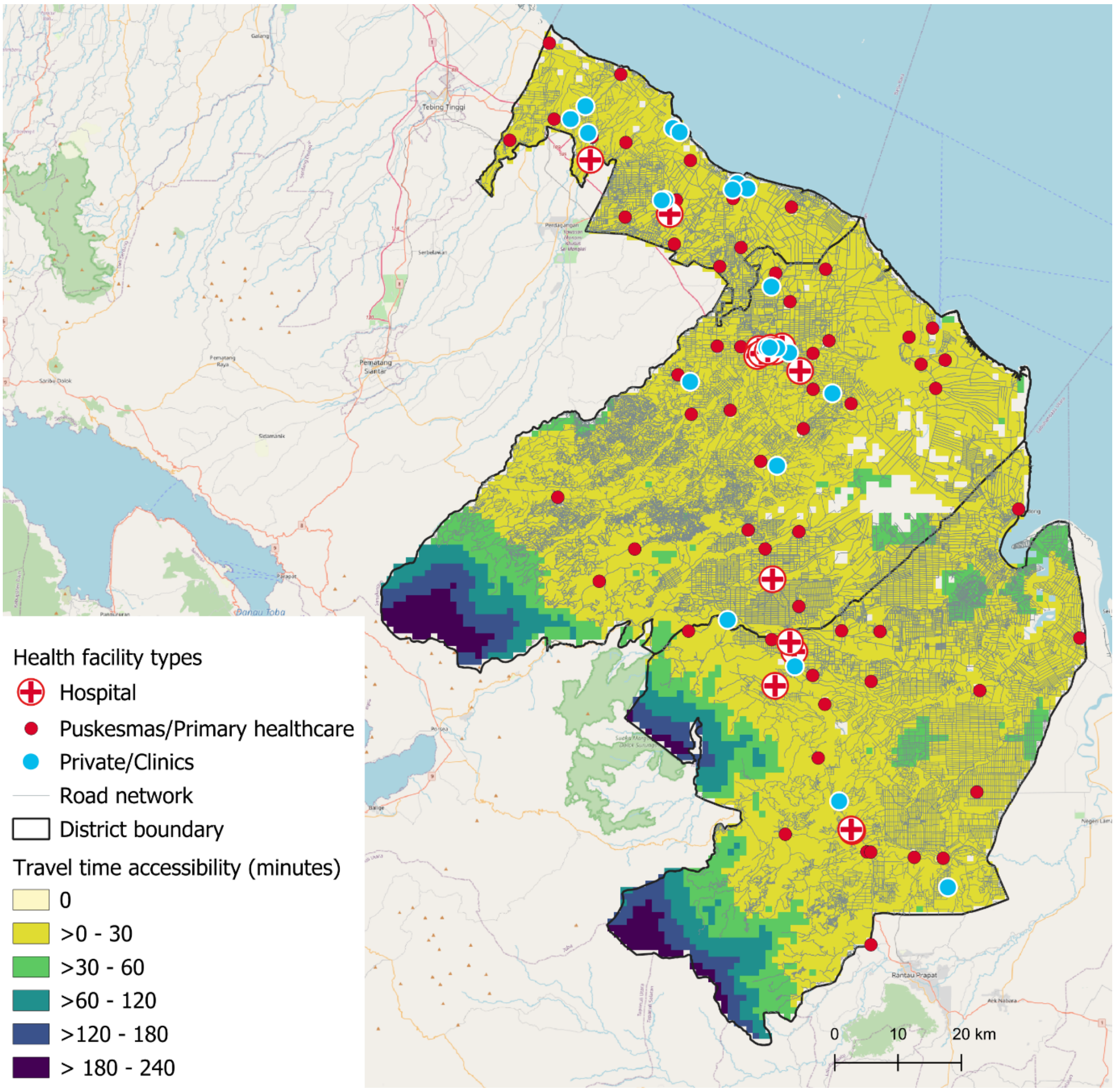

*Figure S2 Accessibility surface from AccessMod with overlaid road network in North Sumatra.*

We customised the estimation of sample referral distances between health facilities using AccessMod to improve accuracy, replacing the simpler default distances assumed by OptiDx. The relationship between the two methodological approaches is summarised in Figure S3.

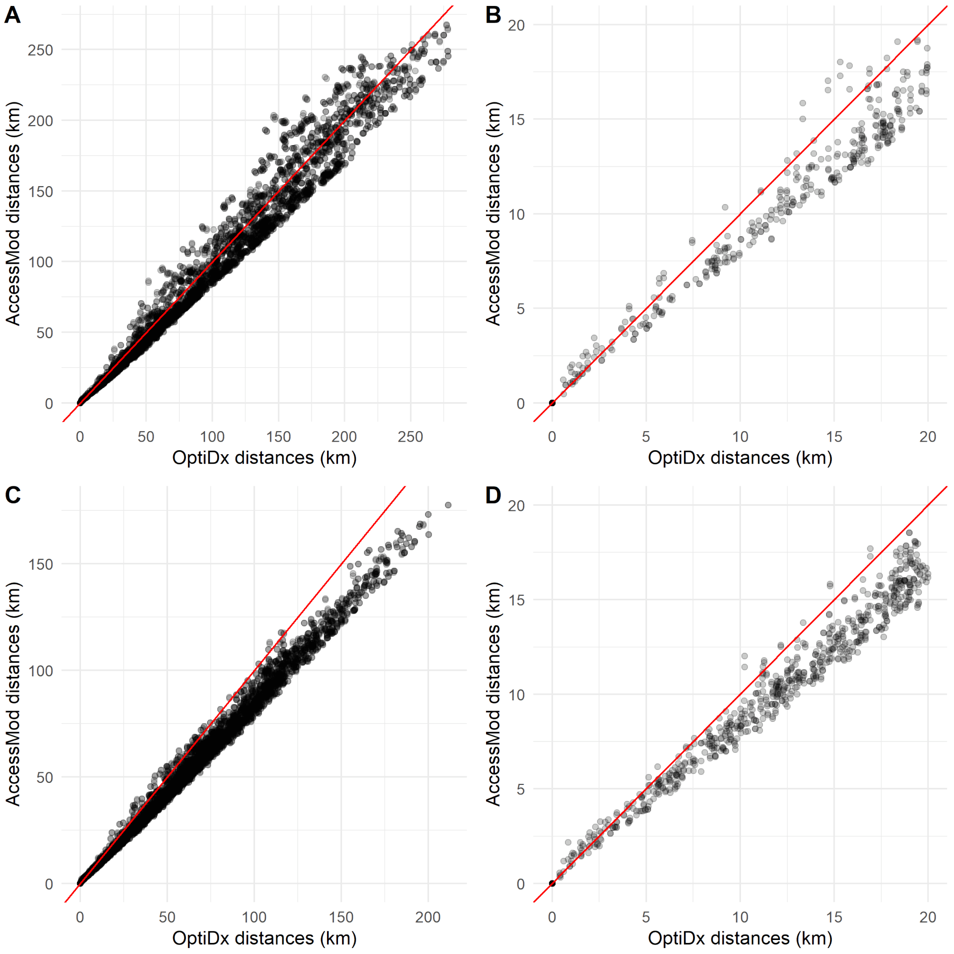

*Figure S3 Scatter plots of the relationship between referral distances (one-way) estimated by AccessMod (y-axis) and OptiDx (x-axis) in (A and B) Sumba and (C and D) North Sumatra. Panels B and D are zoomed-in plots of A and C, respectively, highlighting distances of 20 km or less. The 45-degree diagonal red line represents the equality line, where x = y.*

Distance estimates using OptiDx were generally longer than those from AccessMod, likely leading to consistently higher transportation costs if applied to the models. However, since we restricted healthcare staff travel to 15 km (30 km return trip), the most relevant comparisons are shown in Figures 7C and 7D. Among these locations, the differences were not considered substantial, so the model results should be relatively unaffected by the chosen method. Since AccessMod was assumed to provide greater accuracy, we selected it for our subsequent analysis.
